# Understanding Human–AI Discrepancy in Breast Cancer TIL Assessment: A Multi-Rater and Perceptual Bias Study

**DOI:** 10.64898/2026.05.29.26354196

**Authors:** Abdulkerim Capar, Ibrahim Aloglu, Fugen Aker, Mucahit Ertano, Yusuf Emir Mese, Alptug Ungor, Berfin Ekin Yıldız

## Abstract

**Objective:** Tumor-infiltrating lymphocytes (TILs) in breast cancer are one of the most important indicators of the immune response within the tumor microenvironment. They play a particularly significant prognostic and predictive role in triple-negative and HER2-positive subtypes. However, substantial inter-observer variability has been reported in TIL scoring among pathologists, which limits its reliability in clinical practice. The aim of this study was to evaluate the agreement between artificial intelligence (AI) models and pathologists in TIL scoring and to compare this agreement using different statistical approaches, thereby assessing the potential of AI integration into pathology practice.

**Materials and Methods:** Digitized histopathological images of breast cancer cases were included in the study. Tumor regions annotated by pathologists were evaluated for both stromal TIL percentage and the proportion of stromal tumor area within each ROI, with assessments performed independently by three pathologists and two AI models. Agreement was assessed among pathologists, between pathologists and AI, and between AI models. Statistical analyses included intraclass correlation coefficient (ICC), Cohen’s and Fleiss’ kappa, correlation tests, and Bland–Altman analysis. In addition, categorical agreement was examined using different cut-off values.

**Results:** Inter-pathologist agreement was high, with an ICC of 0.81. In contrast, the global agreement between pathologists and AI models was lower (ICC 0.41). Pairwise comparisons of pathologist–AI agreement yielded substantially lower ICC values (0.12–0.21), although this improved to 0.53 when three pathologists were assessed jointly with a single AI model. The strongest categorical agreement was observed with dichotomized TIL scores (≤10% vs. >10%), whereas multi-category classifications were associated with a marked reduction in kappa values. Spearman correlation coefficients between pathologists and AI models ranged from moderate to good (ρ = 0.48–0.81). Agreement between the two AI models themselves was moderate, with an ICC of 0.64.

**Conclusion:** This study demonstrated that AI models were able to show a certain degree of correlation with pathologists in TIL scoring but remained insufficient in terms of absolute agreement. Differences were more pronounced at higher TIL scores. These findings highlight the need for standardized algorithms trained on larger and more diverse datasets before AI-based TIL scoring can be reliably implemented in clinical practice.

## 1. INTRODUCTION

Breast cancer remains the most frequently diagnosed malignancy among women worldwide and represents a leading cause of cancer-related mortality. In recent years, the tumor microenvironment has gained increasing attention as a critical determinant of disease progression and treatment response. Among its components, tumor-infiltrating lymphocytes (TILs) have emerged as important prognostic and predictive biomarkers, particularly in triple-negative and HER2-positive subtypes. High stromal TIL levels have been shown to correlate with favorable outcomes and improved response to chemotherapy and immunotherapy (Loi et al., 2013; Denkert et al., 2018). Accordingly, TIL evaluation holds significant clinical value in predicting response to chemotherapy and immunotherapy.

Although the International TILs Working Group has published guidelines to standardize TIL assessment (Salgado et al., 2015) interobserver variability remains an important challenge in pathology practice (Swisher et al., 2016). Visual estimation of stromal TIL percentage is inherently subjective and can be influenced by factors such as heterogeneity of lymphocyte distribution, ambiguous stromal boundaries, and individual perceptual biases. Similar work has also characterized inter-observer agreement in stromal segmentation and lymphocyte detection tasks, further emphasizing the inherent variability in human assessments (Capar et al., 2024). Such variability limits the clinical utility of TIL scoring and motivates the search for more objective and standardized approaches.

In recent years, artificial intelligence (AI) and deep learning methods have been increasingly adopted in digital pathology. For AI models to be integrated into pathology practice, not only technical accuracy but also reproducibility and agreement with pathologists’ assessments must be demonstrated (Hendry et al., 2017). Breast pathology has become one of the most active subspecialties for AI research, largely due to the high incidence of breast cancer, the observer-dependent nature of diagnostic and prognostic evaluations, and the availability of large-scale digital histopathology datasets. A wide range of AI models have been developed, from classical machine learning algorithms such as support vector machines and random forests to modern architectures including convolutional neural networks (CNNs) and transformers. Their applications can broadly be categorized as diagnostic, predictive, and prognostic (Unger & Kather, 2024). In diagnostic tasks, AI systems have achieved high performance in tumor detection, cancer grading, and lymph node metastasis identification. For example, the CAMELYON16 challenge demonstrated that some deep learning models could achieve near-pathologist accuracy (Ehteshami Bejnordi et al., 2017). Predictive applications have included the identification of hormone receptor status (ER, PR, HER2) and PD-L1 expression directly from H&E slides, as well as prediction of neoadjuvant chemotherapy response through multimodal AI systems integrating whole slide images and clinical factors (Bai et al., 2021; Li et al., 2022; Liu et al., 2024; Makhlouf et al., 2023; Mao et al., 2025; Thagaard et al., 2021; Zeng et al., 2024) Prognostic applications, such as automated TIL quantification, mitotic figure detection, and histological grading, have shown promising results, with some models providing prognostic information comparable to or exceeding that of human experts (Chaurasia et al., 2025; Jaroensri et al., 2022; Matsumoto et al., 2025; Muthu et al., 2025; Pantanowitz et al., 2020; Wang et al., 2022) Moreover, commercial AI tools such as Ibex’s Galen Breast and Paige Breast Suite have been reported to achieve successful results in multiple studies (Lami et al., 2024; Retamero et al., 2024).

Given these challenges, there is a growing need to systematically investigate not only the technical performance of AI models but also the nature of their agreement with pathologists. Most previous studies have focused on model accuracy relative to reference annotations, whereas fewer have directly compared multiple human observers and AI models evaluating the exact same stromal regions (Kos et al., 2020; Makhlouf et al., 2023; Sun et al., 2021; Vidal et al., 2024). Additionally, the cognitive limitations of human visual estimation—particularly in perceiving small, scattered objects—have received limited attention, despite evidence that such perceptual constraints contribute significantly to interobserver variability.

In this context, the present study aimed to comprehensively evaluate the agreement between pathologists and AI-based TIL scoring of breast cancer. By integrating classical interobserver agreement metrics, correlation analyses, and Bland–Altman assessments with a complementary experiment designed to probe human perceptual bias using synthetic images, this work seeks to elucidate the sources of human–AI discrepancy. Understanding these discrepancies is essential for defining realistic expectations for AI-assisted TIL scoring and for determining how such tools can best be incorporated into future pathology workflows.

## 2. MATERIAL AND METHOD

The experimental workflow of the study is illustrated in Figure 1. First, breast cancer H&E whole-slide images were digitized and stromal regions of interest (ROIs) were manually annotated by pathologists. These ROIs were then processed through the AI pipeline, which included stromal segmentation and lymphocyte detection, to predict TIL scores and stromal ratios (see Figure 3). In parallel, pathologists evaluated the same ROIs and recorded stromal TIL percentages as a rater study. All human-and AI-derived scores were compiled into a consolidated dataset and subjected to comprehensive statistical analyses, including inter-observer agreement metrics, correlation assessments, and Bland–Altman comparisons. To further investigate the sources of human–AI discrepancy, a complementary perceptual bias study was performed using synthetic images designed to mimic scattered lymphocyte distributions.

**Figure 1:**
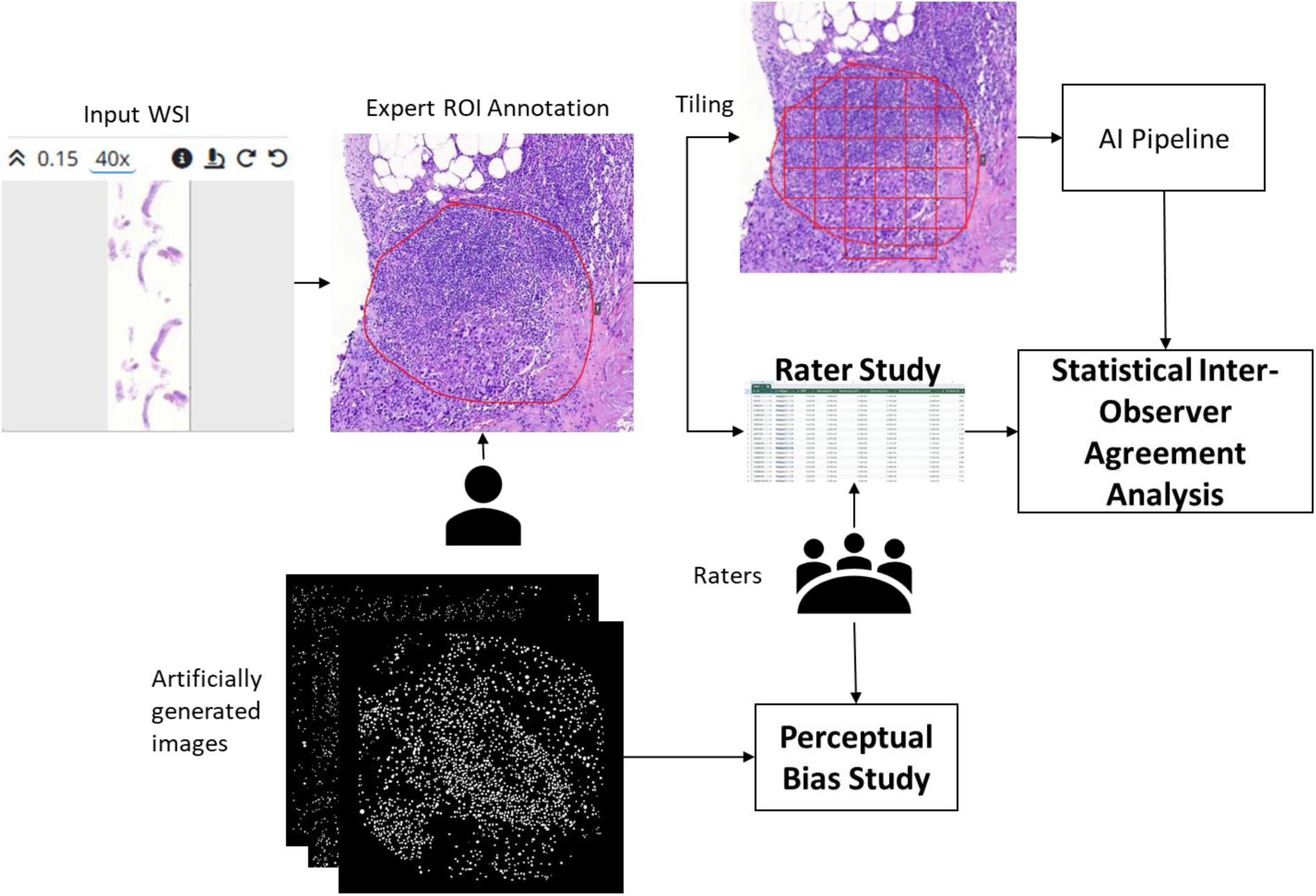
Experimental workflow of the proposed study

### 2.1 Clinical Dataset

This study was approved by the Clinical Research Ethics Committee of the University of Health Sciences Haydarpaşa Numune Training and Research Hospital (approval no. HNEAH-GOAEK/KK/2025/60, dated April 22, 2025). Breast carcinoma cases diagnosed between 2022 and 2024 were retrospectively reviewed from the pathology archives. Among 1,483 available cases (breast-conserving surgery, mastectomy, and tru-cut biopsy specimens), those classified immunohistochemically as triple-negative, HER2-positive, or Luminal B subtype were considered eligible. Cases with adequate tumor areas containing stromal TILs were included, while those lacking archival slides, with insufficient tissue, or with technical artifacts were excluded. A total of 184 cases (68 tru-cut biopsies and 116 excision specimens) met the inclusion criteria and were selected for analysis.

### 2.2 Slide Digitization, Annotation, and Pathologist Scoring

Hematoxylin and eosin (H&E)–stained slides were digitized at ×40 magnification using the MoticEasyScan Pro 6 scanner and uploaded to the EasyPath digital pathology platform (Capar, 2022). A pathologist (I.A.) manually annotated 416 stromal regions within invasive tumor areas, ensuring exclusion of necrosis, crush artifacts, and non-representative fields (Figure 2). These manually defined regions of interest (ROIs) were subsequently evaluated independently by three observers—one expert breast pathologist, one senior resident, and one junior resident. For each ROI, the observers estimated (i) the stromal TIL percentage according to the International TILs Working Group guidelines (Salgado et al., 2015) and (ii) the proportion of stromal tumor area within the total ROI area. All individual numeric assessments were recorded in a structured database for subsequent statistical analyses, including inter-observer agreement measurements, correlation analyses, and comparison with AI-derived scores.

**Figure 2:**
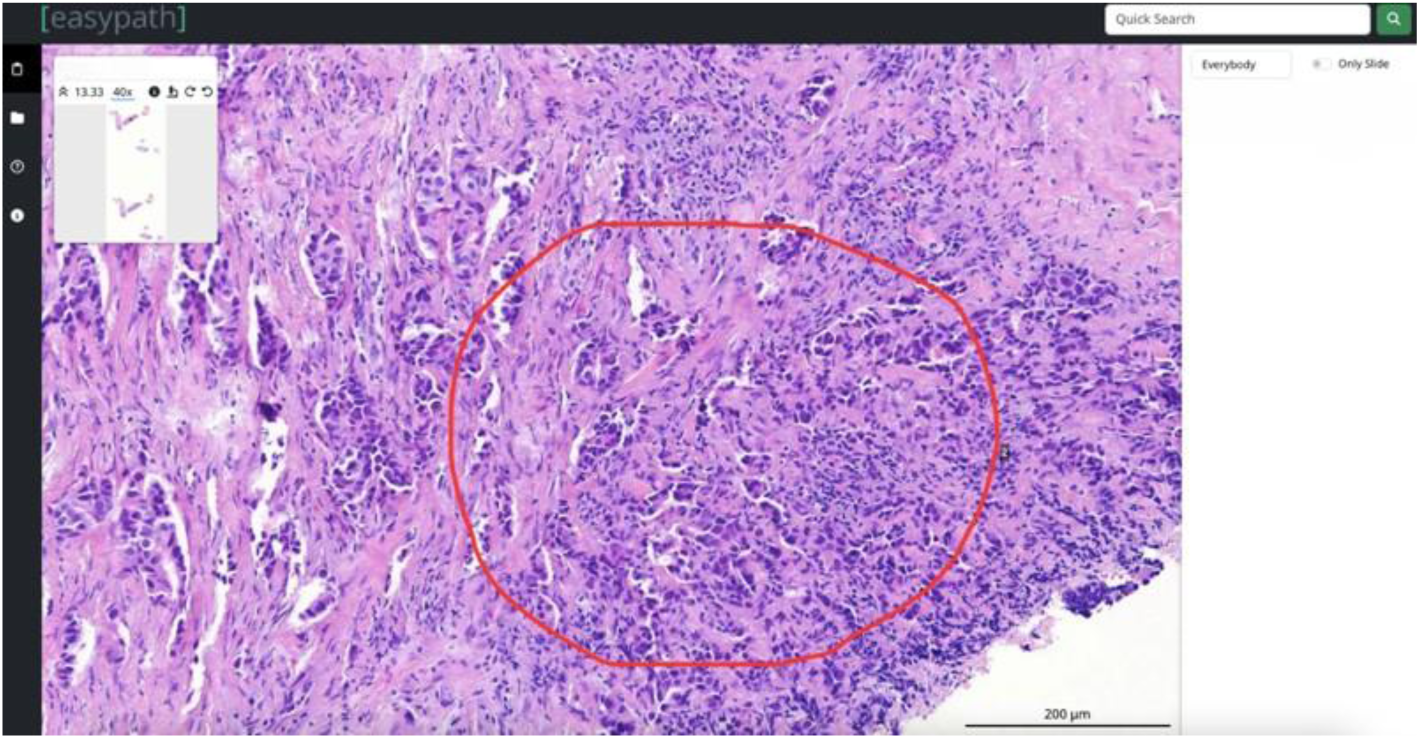
Annotation on the Easypath Platform

**Figure 3:**
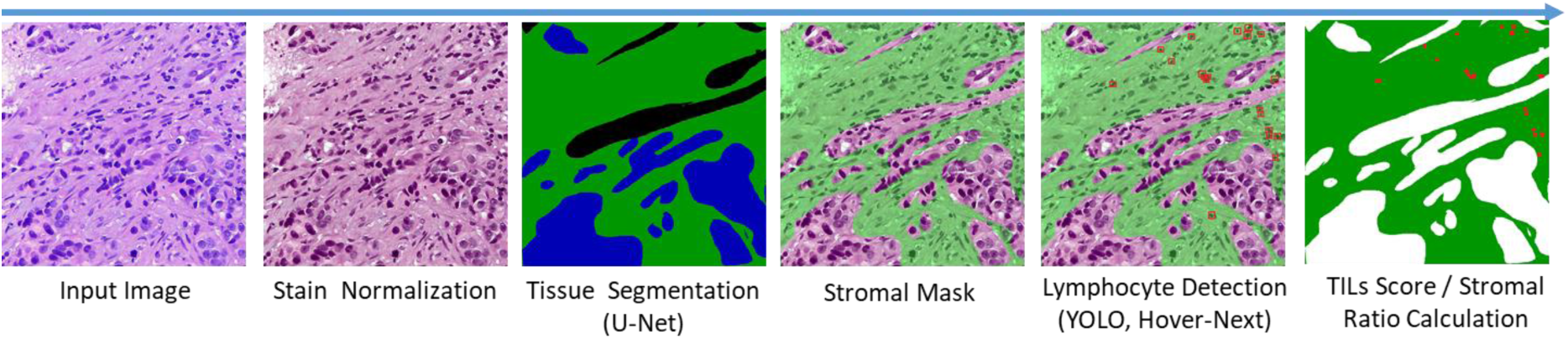
AI based TILs scoring pipeline

### 2.3 AI Based TIL Scoring

The workflow of the proposed AI-based TILs scoring system is illustrated in Figure 3. The procedural stages of the proposed system encompass stain normalization, stromal segmentation utilizing a semantic segmentation deep learning model (UNet), stromal masking applied to stain-normalized input images, lymphocyte detection facilitated by either an object detection (YOLO) or instance segmentation deep learning model (HoVer-NeXt), and ultimately, TIL score computation. Two separate AI pipelines were implemented in this study: YOLO, which combines U-Net–based stromal segmentation with YOLO-based lymphocyte detection, and HoVerNeXt, which integrates the same stromal segmentation step with the HoVer-NeXt instance segmentation model. In addition to TIL quantification, the AI system also calculates the stromal ratio, defined as the proportion of stromal tissue area within the entire ROI.

Deep learning models in the study are trained solely on publicly available datasets without incorporating any local cases. This design ensured that the AI models were evaluated in a completely unbiased manner, allowing their clinical alignment with pathologists to be assessed without any potential benefit from domain-specific fine-tuning.

### Stromal Segmentation Methods

For stromal area segmentation, U-Net based models trained on publicly available research datasets were employed. UNet-based models (Ronneberger et al., 2015) with different kernels are utilized to segment stromal regions. Pytorch Python code library is used to train the models with kernels ResNet, MobileNet, ResNext, and EfficientNet. UNet models are trained and validated on the BCSS (Amgad et al., 2019) and Tiger Challenge (Diagnostic Image Analysis Group of the Radboud University Medical Center, n.d.) datasets.

Histopathological images, especially those derived from different laboratories or staining protocols, often exhibit variations in color, intensity, and brightness due to differences in tissue preparation techniques and staining procedures. These variations can significantly impact the performance of AI models, leading to inaccurate TILs scoring results. Stain normalization methods aim to standardize the appearance of histopathological images by mitigating these color discrepancies, ensuring that the AI models can effectively learn and generalize from the data. In this study, we employed the Reinhard stain normalization technique (Reinhard et al., 2001) to address color inconsistencies in histopathological images.

### Lymphocyte Detection Methods

Lymphocyte detection was performed using two deep learning models—YOLO and HoVer-NeXt—each providing instance-level or pixel-level identification of lymphocytes and plasma cells.

You Only Look Once (YOLO) is a cutting-edge, state-of-the-art family of computer vision models. It is a neural network that predicts the position of bounding rectangles and classification probabilities for an image in one pass. It stands out as one of the widely employed object detection models, recognized for its exceptional speed and accuracy. The first version of YOLO model was proposed by Redmon et al. in 2016 (Redmon et al., 2016). YOLO-v8 model is selected for lymohocyte detection and trained on publicly available datasets PanNuke (Gamper et al., 2020) and NuCLS (Amgad et al., 2022).

HoVer-NeXt is an advanced deep learning architecture designed for nuclear instance segmentation in histopathology images (Graham et al., 2024). Unlike bounding-box–based detectors, HoVer-NeXt predicts horizontal and vertical distance (HoVer) maps together with semantic segmentation outputs, enabling accurate delineation of individual nuclei and effective separation of touching or overlapping lymphocytes. This pixel-level instance segmentation strategy offers a significant advantage in dense inflammatory regions where traditional detection models often fail. HoVer-NeXt builds upon the HoVer-Net framework originally proposed by Graham et al. (Graham et al., 2019), which introduced the use of HoVer maps for robust nucleus separation. In this study, HoVer-NeXt was trained exclusively on publicly available datasets, including PanNuke (Gamper et al., 2020) and the CoNIC challenge dataset (Graham et al., 2024), without incorporating any local cases. This ensured that the model’s performance and clinical agreement with pathologists were evaluated in a fully unbiased manner, independent of domain-specific fine-tuning.

### 2.4 Statistical Inter-Observer Agreement Analysis

All statistical analyses were performed using SPSS version 26 and Jamovi version 2.7. Interobserver and pathologist–AI agreement was quantified using intraclass correlation coefficients (ICC, two-way mixed-effects model, ICC (3,1)), considering both consistency and absolute agreement. Bootstrap resampling (2,000 iterations) was used to calculate 95% confidence intervals when normality assumptions were not met. Additional analyses included Pearson and Spearman correlation coefficients, concordance correlation coefficient (CCC), and kappa statistics. Pairwise Cohen’s kappa values were computed for each observer pair, while Fleiss’ kappa quantified overall agreement among the five raters (three pathologists and two AI models). Agreement levels were interpreted according to conventional thresholds. Bland–Altman plots, scatter plots with regression lines, and boxplots were generated to visualize differences and agreement patterns. A p-value <0.05 was considered statistically significant.

### 2.5 Perceptual Bias Study

To evaluate human perceptual limitations in estimating scattered cellular patterns, we generated synthetic black-and-white images that mimic lymphocyte distributions with controlled levels of clustering (Figure 4). Each image was initialized as a black background, and a set of cluster centers was first placed randomly across the image. Around each center, a group of white circular objects was generated, with their positions sampled from a two-dimensional Gaussian distribution. This produced visually distinct aggregates of small round shapes, resembling lymphocyte clusters frequently observed in inflammatory stromal regions. The number of circles assigned to each cluster was either kept constant or allowed to vary slightly to ensure natural-looking variability. Circle radii were sampled from a small uniform range (e.g., 2–5 pixels) to reflect the approximate size variation of lymphocytes. Overlapping between circles was permitted, allowing dense regions to form and enhancing the realism of the synthetic patterns.

**Figure 4:**
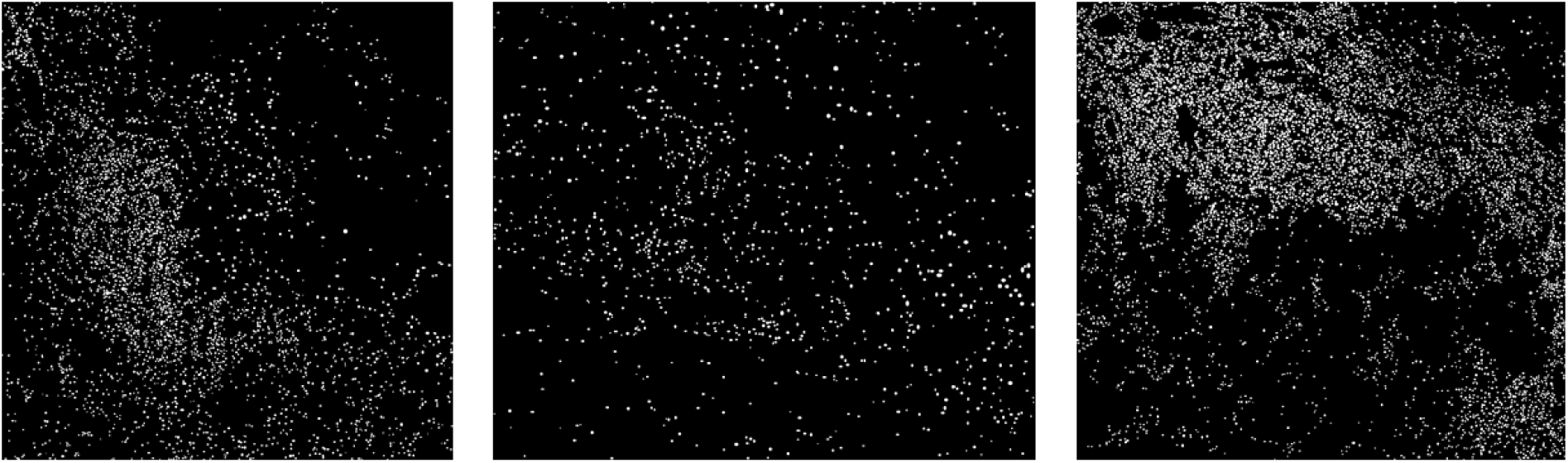
Sample synthetic images generated for perceptual bias study with white region ratios 5.4 %, 2.3 % and 10.8 % from left to right.

To meet a predefined white-area percentage (e.g., 5%, 10%, 20%), the total white-pixel count was monitored during generation, and additional circles were added until the target area proportion was reached within a small tolerance. This approach produced standardized images with the same total white-area ratio but different spatial organizations—clustered versus more uniformly scattered—enabling a focused analysis of how spatial structure influences human estimation and contributes to perceptual bias.

Ten artificially generated black-and-white images were evaluated by 10 pathologists with varying levels of experience (3 months–35 years). Observers estimated the percentage of white area in each image. Agreement between observer estimates and reference values was assessed using ICC.

### 2.6 Hyperparameter Configuration and Computational Environment

For the YOLO training process, the model was trained for 300 epochs using 256×256 image size. Additional training parameters, including optimizer settings and learning rate scheduling, were configured to ensure stable convergence. The training was performed using an NVIDIA GeForce GTX 1080 Ti GPU.

For the segmentation task, training was conducted using a learning rate of 0.01 with a batch size of 32. The model was optimized using the Adam optimizer with the mean squared error (MSE) loss function. An ExponentialLR learning rate scheduler with a decay factor of γ = 0.98 was applied to facilitate stable convergence. The encoder backbone was initialized with ImageNet pretrained weights. Segmentation experiments were executed on an NVIDIA GeForce GTX 1080 Ti GPU under identical software configurations to ensure reproducibility.

The HoVer-Next model was utilized with pretrained weights and was not subjected to any training or fine-tuning. All experiments were conducted in a consistent computational environment to ensure reproducibility.

## 3. RESULTS

### Cohort characteristics

A total of 186 cases from 179 patients were included. The mean age was 55.5 ± 13.3 years and the median was 54 (range 29–96) years; 99.4% (n=178) were female. Specimen types were tru-cut biopsies (66.3%, n=118), breast-conserving surgery (19.7%, n=35), and mastectomy (14.0%, n=25); one case was a subcutaneous mastectomy (0.6%). Histologic subtypes were predominantly invasive ductal carcinoma (94.4%, n=170), with rare invasive lobular carcinoma (1.1%, n=2) and mixed IDC+ILC (1.1%, n=2). Molecular subtypes were Luminal B (44.4%, n=79), triple-positive (21.9%, n=39), triple-negative (19.7%, n=35), and HER2-positive (14.6%, n=26). Nodal status was pN0 in 18.3% (n=40), ≥pN1 in 9.6% (n=21), and unavailable in 72.1% (n=118) (Table 1). Across 416 ROIs, the total annotated area was 361.92 mm²; the mean ROI size was 0.87 mm² (range 0.11–6.52 mm²).

**Table 1:** Histopathological and Demographic Characteristics.

| Characteristic | n | % |
| --- | --- | --- |
| **Age (years)** |  |  |
| Mean ± SD | 55.5 ± 13.3 | - |
| Median (min–max) | 54 (29–96) | - |
| **Sex** |  |  |
| Female | 178 | 99.4 |
| Male | 1 | 0.6 |
| **Procedure** |  |  |
| Tru-cut | 118 | 66.3 |
| Breast-Conserving Surgery | 35 | 19.7 |
| Mastectomy | 25 | 14.0 |
| Subcutaneous Mastectomy | 1 | 0.6 |
| **Histologic Type** |  |  |
| Invasive Ductal Carcinoma (IDC) | 170 | 94.4 |
| Invasive Lobular Carcinoma (ILC) | 2 | 1.1 |
| IDC + ILC | 2 | 1.1 |
| Other | 5 | 2.8 |
| **Molecular Subtype (Immunoprofile)** |  |  |
| Luminal B | 79 | 44.4 |
| HER2 Positive | 26 | 14.6 |
| Triple Negative | 35 | 19.7 |
| Triple Positive | 39 | 21.9 |
| **Lymph Node Status** |  |  |
| pN0 | 40 | 18.3 |
| ≥pN1 | 21 | 9.6 |
| Unknown | 118 | 72.1 |

### Inter-pathologist TIL score agreements

The global inter-pathologist ICC for TIL percentage was 0.808 (95% CI **0**.76–0.84), with consistency ICC 0.843 (95% CI 0.81–0.87) (Table 2). Pairwise ICCs indicated strong agreement: Path1–Path2 0.82 (0.77–0.86), Path1–Path3 0.80 (0.75–0.85), and Path2–Path3 0.80 (0.75–0.85) (Figure 5). Concordance correlation coefficients (CCC) were concordant with ICCs—0.82, 0.80, and 0.80 for the same pairs, respectively. Spearman correlations were high: ρ=0.82, 0.85, and 0.83 (all p<0.001) (Figure 6), mirrored by Pearson correlations 0.82–0.84 (all p<0.001)

**Figure 5:**
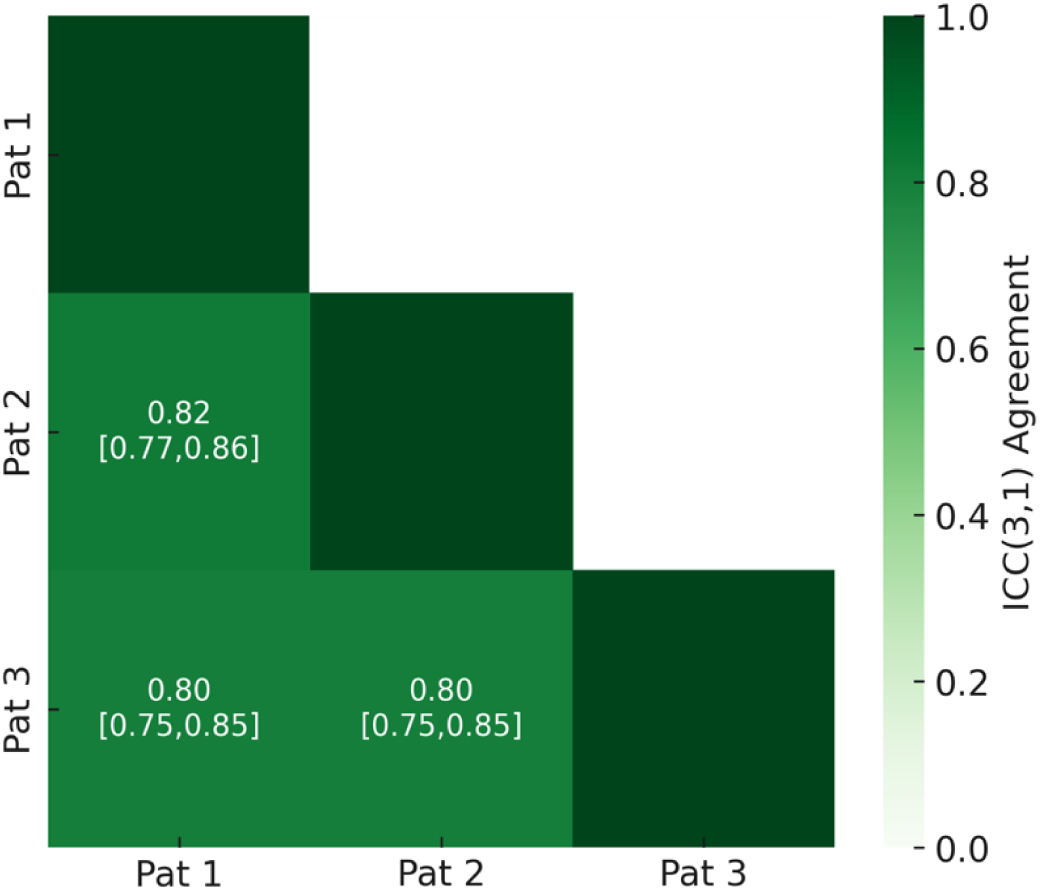
Inter-Pathologist Intraclass Correlation Coefficients (ICC)

**Figure 6:**
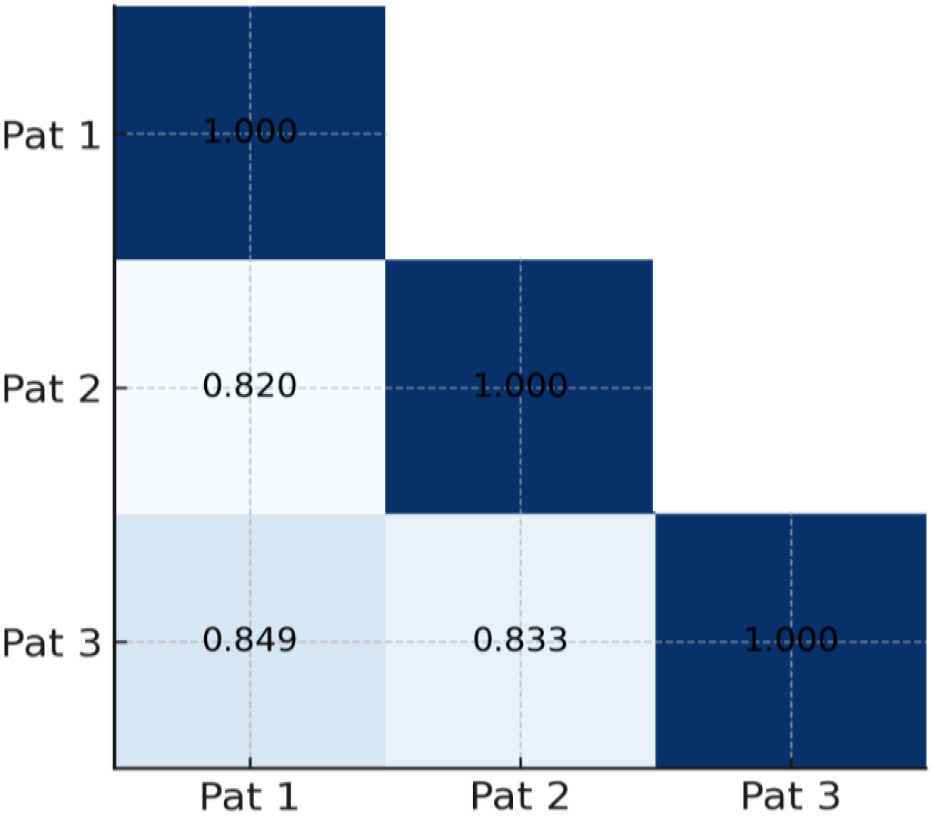
Inter-Pathologist Spearman Correlation Coefficients

**Table 2:** Overall Agreement Among Pathologists.

|  | ROI | Rater | Consistency | Absolute Agreement |
| --- | --- | --- | --- | --- |
| Value | 416 | 3 | 0.843 (0.81-0.87) | 0.808 (0.76-0.84) |

### Categorical inter-pathologist TIL score agreement

Dichotomizing TIL at ≤10% vs >10% yielded Fleiss’ kappa 0.58 (0.52–0.64) and pairwise Cohen’s kappa 0.54–0.67 (0.47–0.74). With three categories (≤10%, 11–30%, ≥30%), Fleiss’ kappa 0.50 (0.45–0.55); pairwise 0.46–0.61 (0.39–0.67). Using ≤10%, 11–40%, ≥40%, Fleiss’ kappa 0.52 (0.46–0.57); pairwise 0.48–0.60 (0.41–0.66) (Table 3). Kappa decreased modestly with more categories, but remained moderate across thresholds.

**Table 3:**
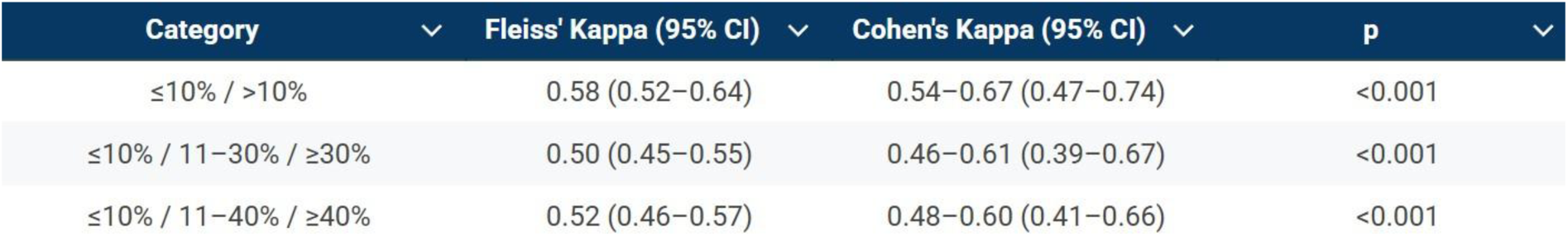
Inter-Pathologist Categorical TIL Score Agreement.

### Agreement between AI models

The two AI models showed moderate agreement: ICC 0.64 (0.54–0.74). Their Spearman correlation was ρ=0.691 (p<0.001).

### Global agreement among all raters (3 pathologists + 2 AI)

Considering five raters, the ICC was 0.41 (0.38–0.43). Grouping 3 pathologists + HoVer-Next yielded ICC 0.53, whereas 3 pathologists + YOLO yielded ICC 0.50 (Table 4).

**Table 4:** Agreement Between Pathologists and Artificial Intelligence Models.

| Group | ICC (3,1) |
| --- | --- |
| 3 Pathologists + HoVerNext | 0.53 |
| 3 Pathologists + YOLO | 0.50 |

### Pairwise agreement: pathologist vs AI

Pairwise ICCs (agreement) between individual pathologists and AI were low (Table 5):

- Path1–HoVer-Next 0.21; Path1–YOLO 0.12
- Path2–HoVer-Next 0.21; Path2–YOLO 0.16
- Path3–HoVer-Next 0.15; Path3–YOLO 0.10

**Table 5:** Pairwise Agreement Between Pathologists and AI Models.

|  | Pat1 | Pat2 | Pat3 |
| --- | --- | --- | --- |
| HoVerNext | 0.21 | 0.21 | 0.15 |
| YOLO | 0.12 | 0.16 | 0.10 |

CCC values closely tracked ICCs (Table 6).

**Table 6:** Inter-Observer Concordance Correlation Coefficients (CCC)

|  | Pat 1 | Pat 2 | Pat 3 | AI 1 | AI 2 |
| --- | --- | --- | --- | --- | --- |
| Pat 1 | 1.00 |  |  |  |  |
| Pat 2 | 0.82 (0.77-0.86) | 1.00 |  |  |  |
| Pat 3 | 0.80 (0.76-0.84) | 0.80 (0.75-0.84) | 1.00 |  |  |
| AI 1 | 0.21 (0.18-0.24) | 0.21 (0.17-0.24) | 0.15 (0.13-0.17) | 1.00 |  |
| AI 2 | 0.13 (0.10-0.16) | 0.16 (0.12-0.20) | 0.10 (0.08-0.13) | 0.64 (0.52-0.74) | 1.00 |

### Correlation: pathologist vs AI

Spearman correlations showed higher associations with HoVer-Next than YOLO (Figure 7):

- Path1–HoVer-Next ρ=0.745, Path1–YOLO ρ=0.482 (both p<0.001)
- Path2–HoVer-Next ρ=0.708, Path2–YOLO ρ=0.503 (both p<0.001)
- Path3–HoVer-Next ρ=0.719, Path3–YOLO ρ=0.529 (both p<0.001) Pearson analyses showed similar trends.

**Figure 7:**
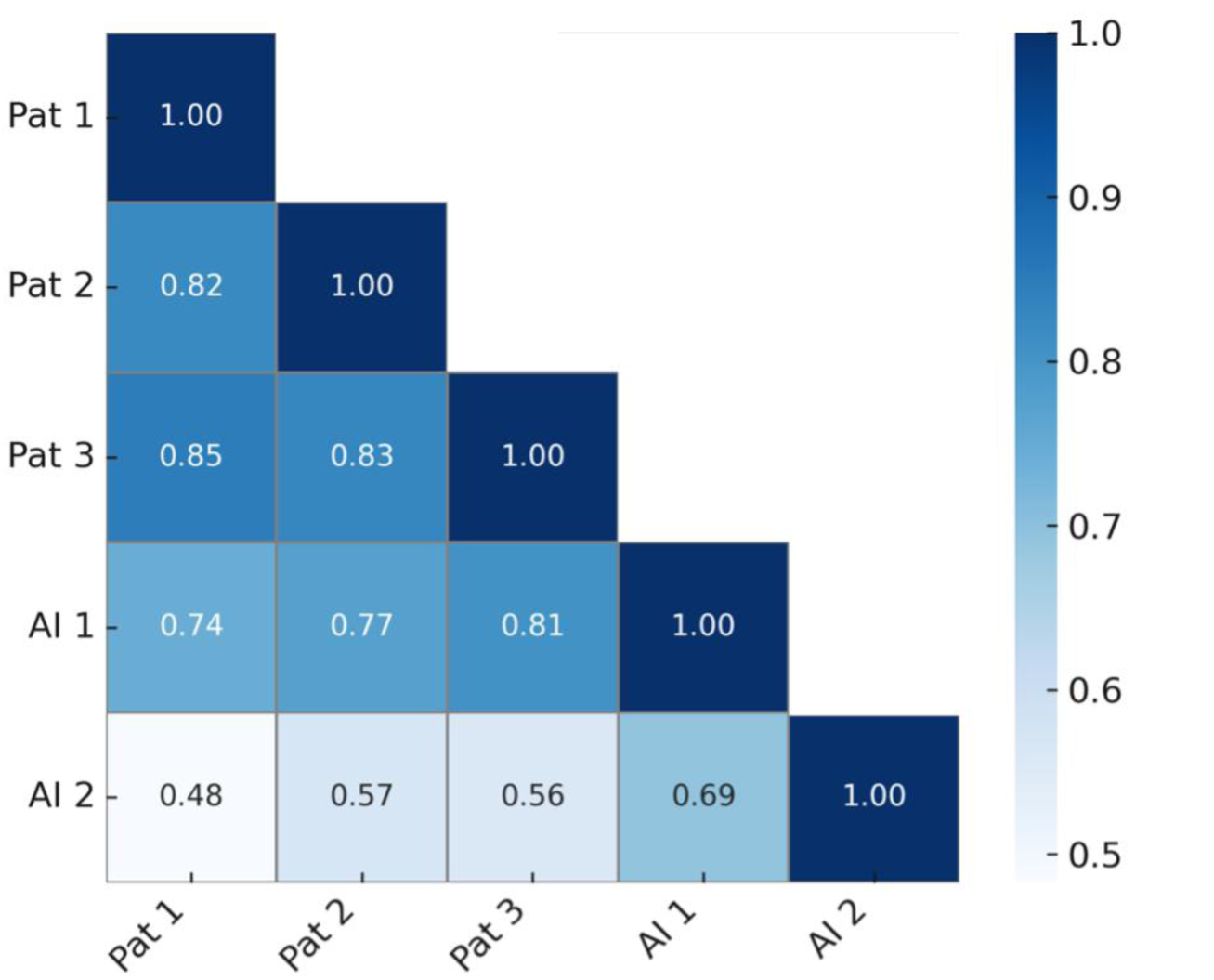
Inter-Observer Spearman Correlation Coefficients

### Categorical agreement: pathologists vs AI

Across cutoffs, overall agreement was low. With ≤10% vs >10%, Fleiss’ kappa 0.30 (0.25–0.35) and pairwise Cohen’s kappa 0.11–0.31 (0.07–0.11). With three categories, agreement further declined (Fleiss’ kappa 0.21–0.23; pairwise 0.02–0.13, 95% CI 0.00–0.19) (Table 7).

**Table 7:** Kappa Values Between Pathologists and AI Models.

| Category | Fleiss' Kappa | Cohen's Kappa | AI–AI Kappa | p |
| --- | --- | --- | --- | --- |
| $\leq 10\%$ / $> 10\%$ | 0.30 (0.25–0.35) | 0.11–0.31 (0.07–0.37) | 0.45 (0.31–0.58) | <0.001 |
| $\leq 10\%$ / 11–30% / $\geq 30\%$ | 0.21 (0.19–0.25) | 0.02–0.13 (0.00–0.19) | 0.45 (0.28–0.56) | <0.001 |
| $\leq 10\%$ / 11–40% / $\geq 40\%$ | 0.23 (0.20–0.27) | 0.02–0.16 (0.05–0.22) | 0.45 (0.33–0.58) | <0.001 |

### Trend plots and bias

Boxplots showed a monotonic increase in AI scores across pathologist-defined categories, but with lower absolute values for AI: pathologist mean 0–10% → AI mean 2.6Figure 8(Figure 8). Scatter plots with regression lines demonstrated a positive but limited linear trend Figure 9).

**Figure 8:**
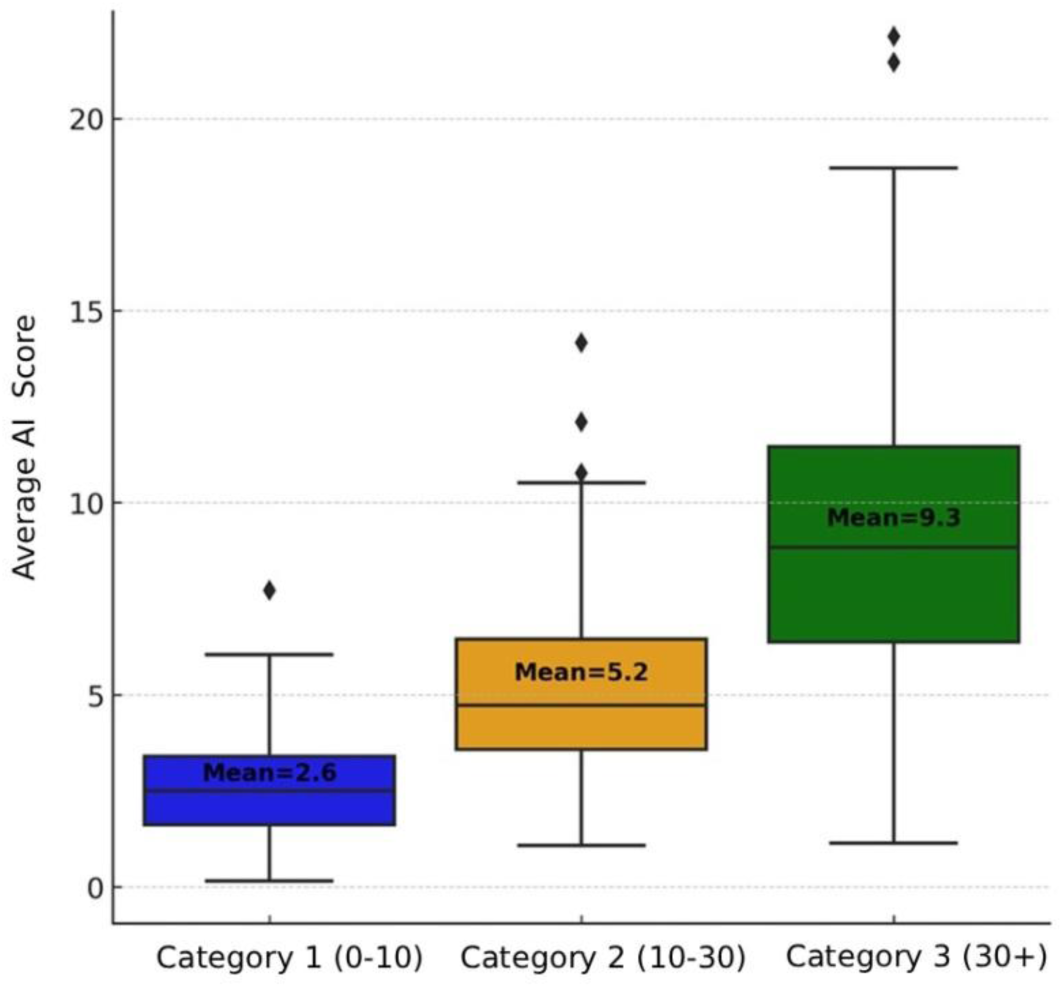
Distribution of Pathologist Averages and Artificial Intelligence Averages Across Different Levels

**Figure 9:**
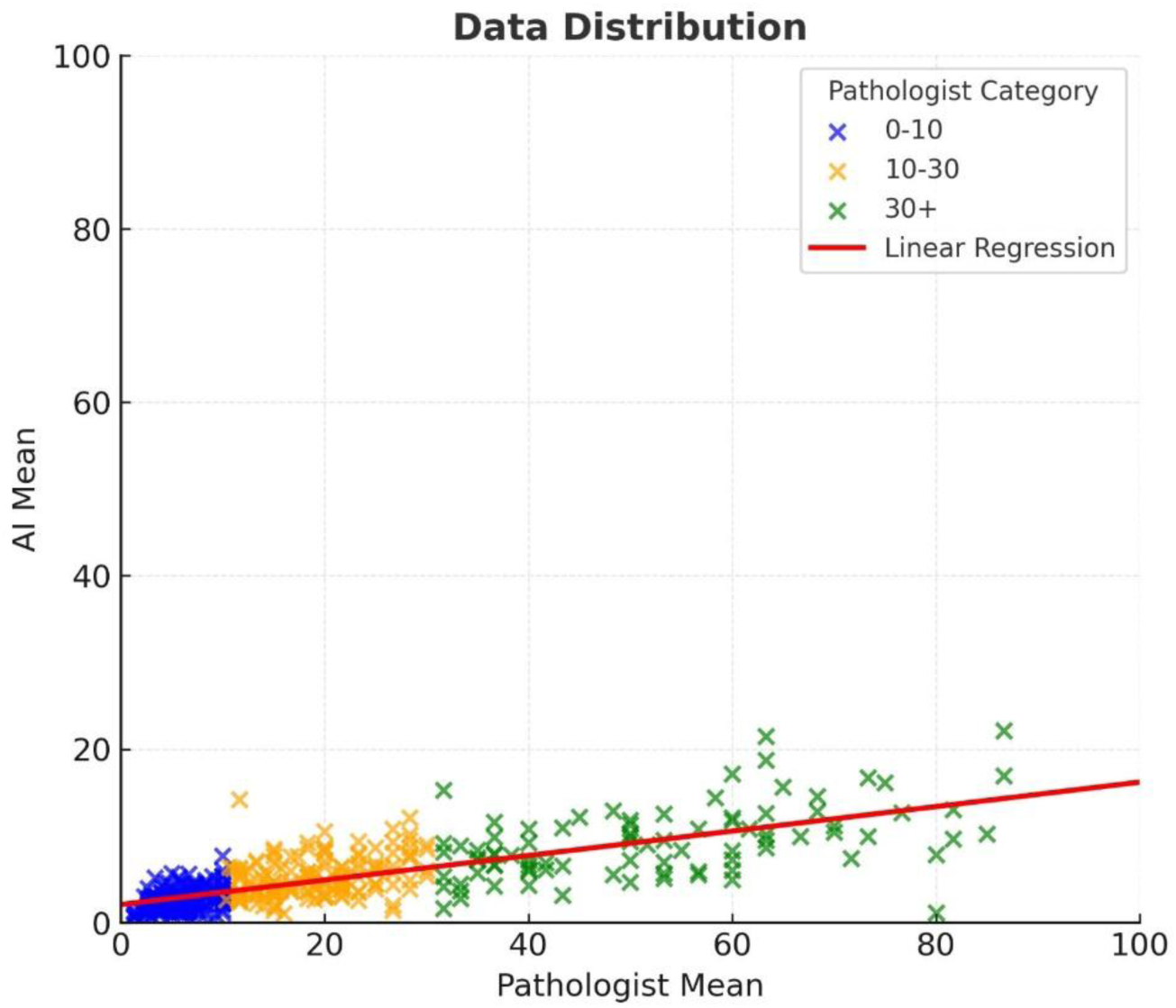
Linear Relationship Between the Pathologist Mean and AI Model Mean

Bland–Altman analyses revealed systematic negative bias for AI vs the mean of pathologists:

- YOLO bias −16.17 (AI lower); limits of agreement −17.67 to 50.00
- HoVer-Next bias −14.90; limits −17.01 to 46.74

Bias and dispersion increased at higher average TIL values (Figure 9 and Figure 10).

**Figure 10:**
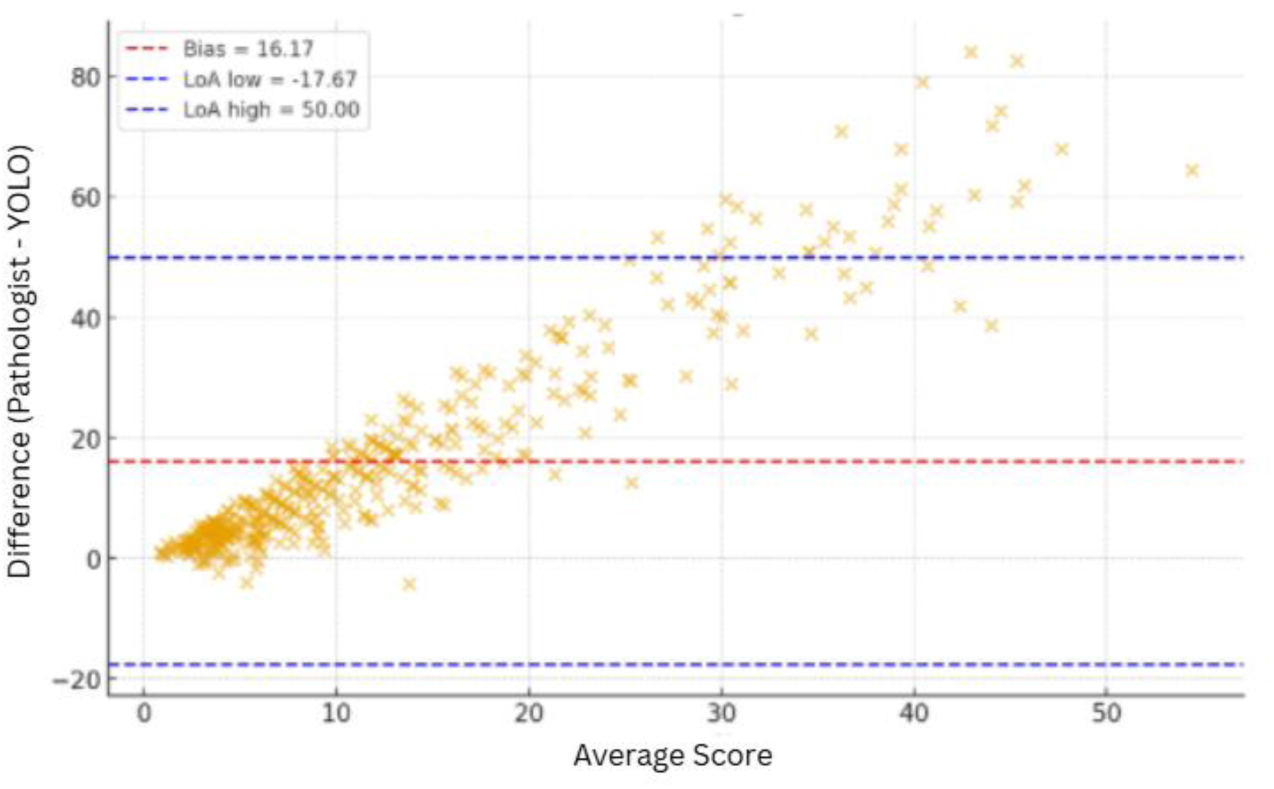
YOLO Model vs. Pathologist Mean

**Figure 11:**
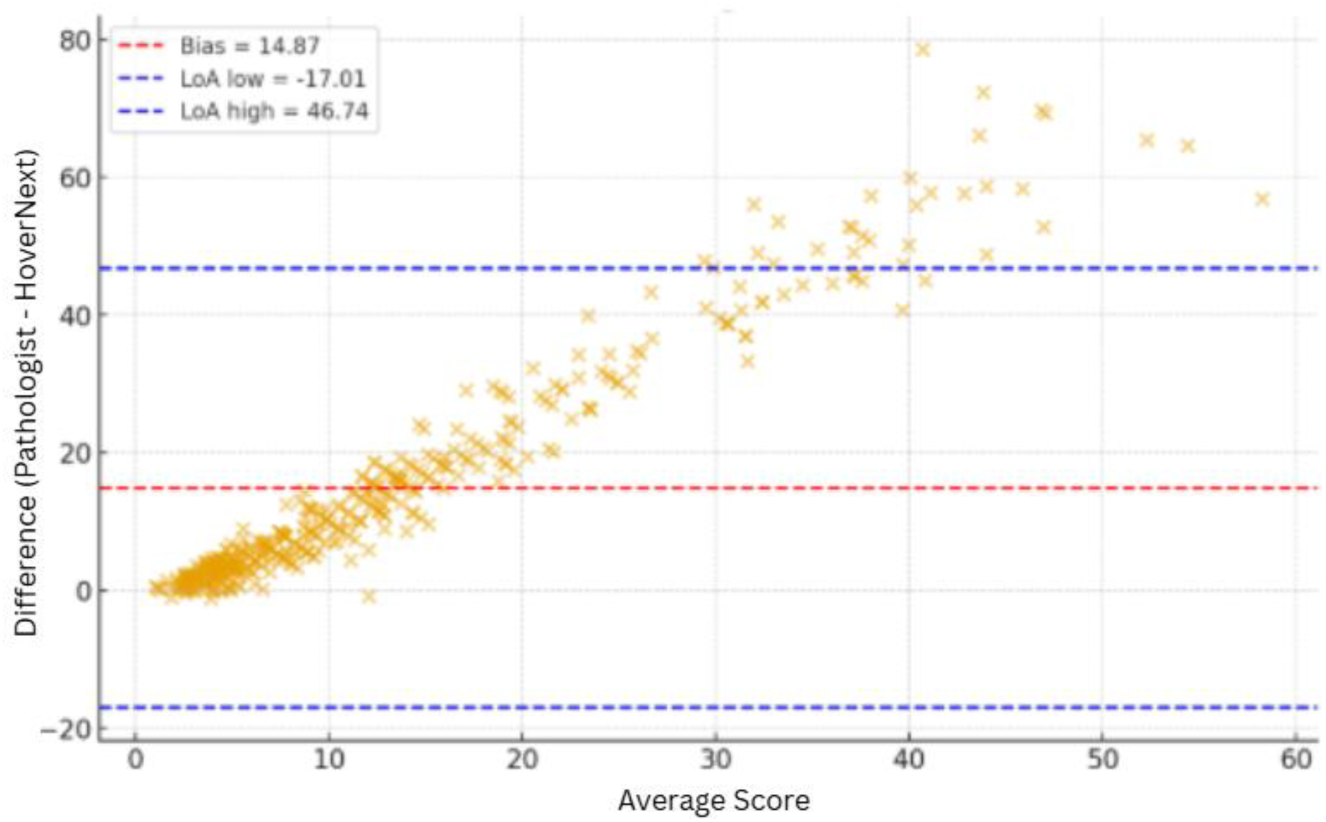
HoVer-NeXt Model vs. Pathologist Mean

### Stromal ratio agreement

Agreement among three pathologists plus the U-Net stromal segmentation model yielded ICC 0.65 (0.60–0.69). The pathologist-only global ICC on the same task was 0.76 (0.71–0.79), indicating higher human-human consistency but comparable performance of U-Net in this component.

### Perceptual bias study results

In the synthetic black-and-white images, agreement between 10 pathologists and ground-truth white-area proportions showed ICC 0.16–0.57, indicating poor-to-moderate concordance and supporting the notion that visual percentage estimation is error-prone, consistent with the main study’s pathologist–AI discrepancies.

Overall, pathologists exhibited good internal agreement, AI models showed moderate mutual agreement, and pathologist–AI agreement was low, with AI systematically underestimating TIL at mid-to-high ranges—HoVerNext closer to pathologist scores than YOLO.

## 4. DISCUSSION

The evaluation of tumor-infiltrating lymphocytes (TILs) has gained increasing clinical importance in breast cancer, serving as both a prognostic and predictive biomarker. In particular, high stromal TIL densities have been associated with improved treatment response and survival in triple-negative breast cancer (TNBC) and HER2-positive subtypes (Denkert et al., 2018; Loi et al., 2013). However, TIL assessment in routine pathology practice remains largely subjective (Choi et al., 2023), leading to substantial inter-observer variability despite existing international guidelines. The International TILs Working Group (TIL-WG) introduced a standardized stromal TIL evaluation guideline in 2014, which has improved reproducibility across observers (Salgado et al., 2015). Nevertheless, differences among pathologists persist and continue to pose a significant challenge for the clinical implementation of TIL scoring.

In this study, we observed high agreement among three pathologists, which was reflected in both continuous and categorical measures. The overall ICC (3,1) value for absolute agreement was 0.80 (95% CI: 0.76–0.84), and pairwise ICCs ranged between 0.80 and 0.82, indicating good to excellent reliability. These findings are consistent with previously published studies reporting moderate-to-high inter-observer agreement when standardized TIL-WG criteria are applied (Cabuk et al., 2018; Tramm et al., 2018; Van Bockstal et al., 2021). Complementary analyses using Lin’s concordance correlation coefficient (CCC) yielded similarly high values (0.80–0.82), reinforcing the strong alignment between observers in both accuracy and precision. Moreover, Spearman (ρ = 0.82–0.84, p < 0.001) and Pearson (r = 0.82–0.85, p < 0.001) correlation coefficients were also high, indicating strong monotonic and linear relationships among pathologists’ TIL scores. Categorical analyses showed moderate agreement, with Fleiss kappa values of 0.58 for binary classification (≤10% vs. >10%) and 0.50–0.52 for multi-category classifications (three-tier systems), consistent with previous reports (Swisher et al., 2016; Tramm et al., 2018). Together, these results confirm that experienced pathologists can achieve reproducible TIL assessments.

In contrast, the agreement between pathologists and AI models was notably lower. When three pathologists and two AI models (HoVerNext and YOLO) were analyzed together, the overall ICC dropped to 0.41 for absolute agreement. Pairwise comparisons between individual pathologists and AI models yielded ICC values between 0.10 and 0.21, indicating poor agreement. Among the models, HoVerNext demonstrated better performance than YOLO, with higher ICCs (0.21 vs. 0.10–0.16). Nonetheless, both models consistently underestimated TIL scores, particularly in medium and high TIL categories, as demonstrated by box plots and Bland–Altman analyses. The mean bias was approximately 15% lower than pathologists’ scores, with wider limits of agreement observed for YOLO compared to HoVerNext. Importantly, the ICC values observed between pathologists and AI models in our study were lower than those reported in several previous investigations, where AI approaches evaluated on either pathologist-annotated regions or whole-slide images typically achieved moderate agreement levels with human observers. (Makhlouf et al., 2023; Sun et al., 2021)

When categorical agreement was assessed, concordance between pathologists and AI models was generally weak. Using a binary cutoff (≤10% vs. >10%), kappa values were low (Fleiss κ = 0.30; Cohen’s κ = 0.11–0.31). Agreement decreased further with three-category classifications (Fleiss κ = 0.21–0.23; Cohen’s κ = 0.02–0.16), indicating limited alignment between AI models and pathologists across different thresholds.

Despite these discrepancies in absolute values, the correlation between pathologists and AI remained moderate to high. Consistent with prior studies, we observed a similar pattern of association (Bai et al., 2021; Vidal et al., 2024). This suggests that while AI models can follow the relative ranking of cases to some extent, they fail to match pathologists’ absolute scoring levels.

To better understand the observed discrepancies, the segmentation and cell detection outputs for the annotated regions were re-examined with the involvement of experienced pathologists (Figure 12 and Figure 13). This review revealed that AI models failed in segmentation or cell detection onlysT in a small number of cases (which were not excluded from the analysis), while the majority of regions were technically processed correctly. Notably, even in regions where AI performed well, a systematic difference between pathologists and AI persisted (pathologists consistently assigned higher TIL scores). This suggests that the disagreement cannot be attributed solely to algorithmic limitations but may also reflect inherent perceptual and cognitive biases in human visual estimation. In cases with marked discrepancies between pathologist and AI scores, the pathologists’ assessments were revisited; however, substantial revisions toward AI estimates were not made, ensuring that the observed differences genuinely represented human–AI divergence.

**Figure 12:**
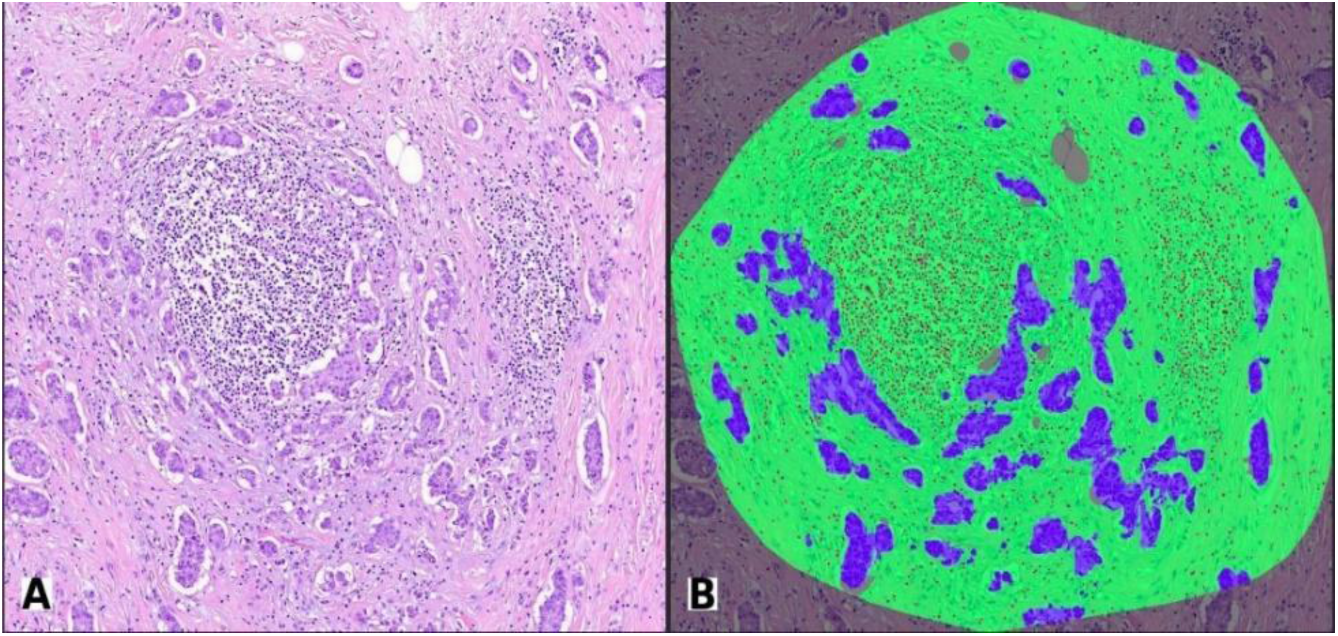
Evaluated Annotation Area A: Annotation Area B: Segmentation and Lymphocyte Detection by the AI Model

**Figure 13:**
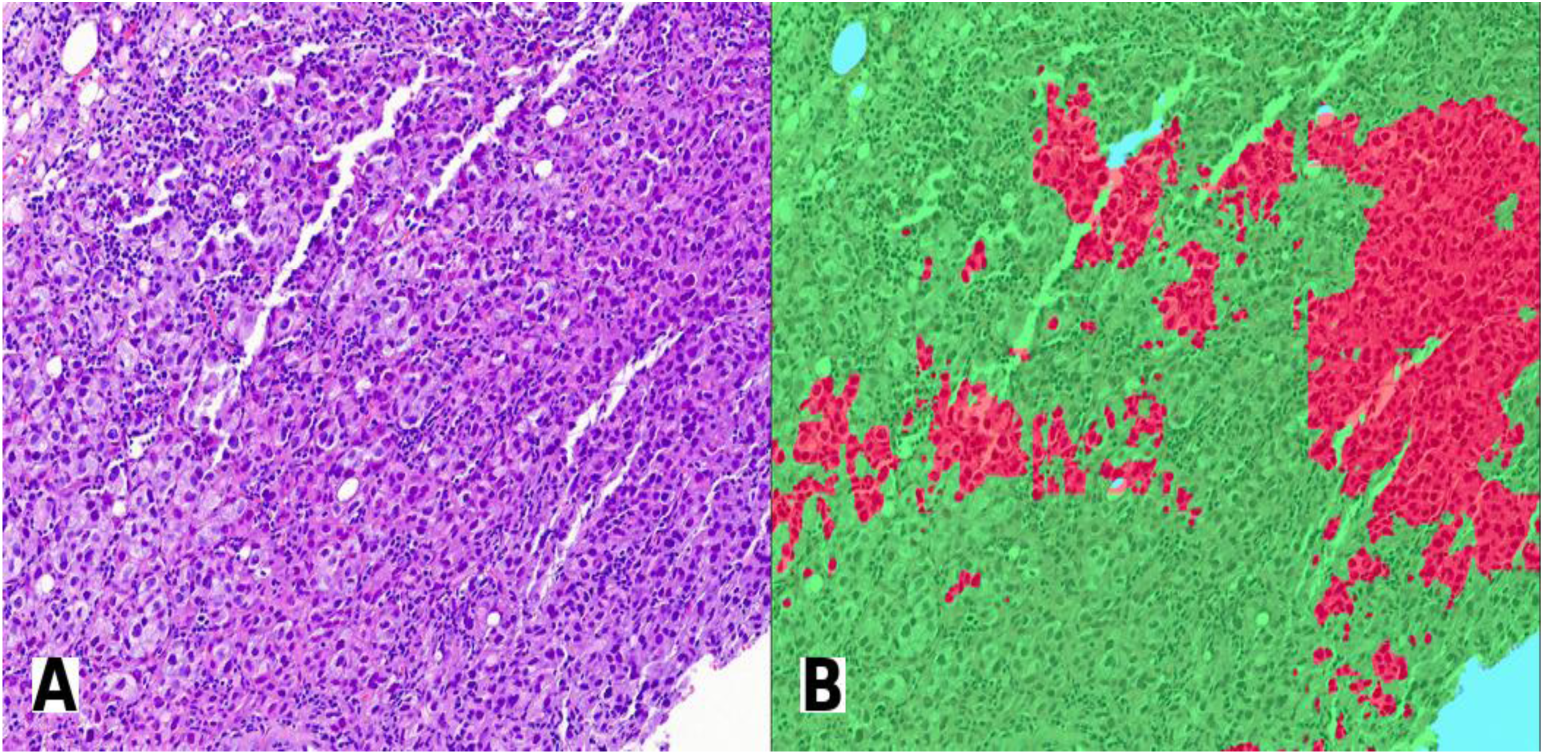
Low-Accuracy Segmentation — A: Original Image, B: Segmentation Output

Building on these observations, we conducted an additional perceptual bias study to specifically test the limitations of human visual estimation in perceiving scattered areas. For this, we designed artificial black-and-white images that mimicked lymphocyte distributions, allowing for exact ground-truth calculations (Figure 4). Pathologists were asked to estimate the proportion of white area in each image, and their scores were compared with the true values. The intraclass correlation coefficients between pathologists and the ground truth ranged from 0.16 to 0.57, indicating weak-to-moderate agreement. These findings closely mirrored the levels of agreement observed between pathologists and AI models in the main study, confirming that human estimation has inherent limitations when dealing with dispersed small objects. Previous work has shown that heterogeneity in lymphocyte distribution and unclear stromal boundaries contribute to interobserver variability (Kos et al., 2020), whereas AI can perform pixel-based segmentation and counting, bypassing these perceptual constraints. Phenomena such as the “crowding effect” and “convex-hull area effect” have been described as key sources of human error in area estimation (Shilat et al., 2021; Yousif & Keil, 2019). Lönnqvist et al. further demonstrated that crowding effects affect human observers much more strongly than neural networks (Lonnqvist et al., 2020). Together, these results support that pathologist–AI discrepancies arise not only from algorithmic factors but also from cognitive limitations, and the use of artificial images provided a robust methodological basis for this conclusion.

This study has several strengths. It involved a large sample size, multiple pathologists, and two different AI models applied to the same dataset, allowing for a comprehensive comparison. Multiple complementary statistical methods (ICC, kappa, CCC, correlation, and Bland–Altman analyses) were employed to ensure robust conclusions. Furthermore, the pivot study provided a unique methodological contribution by demonstrating the cognitive basis of human–AI discrepancies.

Nonetheless, several limitations must be acknowledged. The AI models evaluated here were applied to pathologist-defined tumor ROIs rather than whole-slide images, limiting their autonomy and potentially underestimating their clinical applicability. The models used (HoVerNeXt and YOLO for lymphocyte detection, U-Net for stromal segmentation) were pre-trained networks that were not fine-tuned on our local dataset, which may have impacted performance. However, this design choice also represents a strength of the study: by avoiding local fine-tuning, the evaluation reflects a more realistic clinical scenario in which pre-trained, off-the-shelf models are applied to new institutional data without prior adaptation. This allowed us to assess human–AI agreement under unbiased, domain-shifted conditions and provides insight into the true generalizability of current AI methods. Finally, this was a single-center study, and multicenter validation on larger datasets will be needed to further support and generalize the findings.

## 5. CONCLUSION

In this study, we demonstrated that while inter-pathologist agreement in TIL scoring was consistently high across multiple statistical measures, current AI models still exhibit limited concordance with human observers, particularly in medium and high TIL categories. This discrepancy persisted despite technically accurate segmentation and detection outputs, indicating that differences are driven not only by algorithmic limitations but also by intrinsic perceptual biases in human visual estimation. Supporting this, our complementary pivot study confirmed that pathologists systematically underestimate or overestimate scattered areas, highlighting the cognitive constraints underlying TIL assessment.

These findings underscore both the promise and current limitations of AI-assisted TIL scoring. While AI algorithms already provide objective and reproducible quantification, further optimization, standardization, and validation on large multi-institutional datasets are required before their reliable clinical integration. AI is likely to evolve from a standalone scoring tool into a decision-support mechanism that enhances diagnostic standardization and reduces observer variability in pathology practice. The systematic underestimation observed suggests that future AI systems must incorporate not only improved detection accuracy but also perceptual calibration strategies aligned with human visual tendencies.

## Data Availability

The data that support the findings of this study are not publicly available due to patient privacy and institutional restrictions, but may be available from the corresponding author upon reasonable request and with permission from the relevant institution.

